# Negative pressure pneumomediastinum: a novel concept of spontaneous pneumomediastinum without mediastinal widening following weight loss

**DOI:** 10.1101/2022.08.02.22277892

**Authors:** Hiroaki Hagiwara, Yoshinori Kinno, Tadashi Ikegami

**Author notes:** Corresponding author: (HH).

## Abstract

**Purpose:** Spontaneous pneumomediastinum, supposedly attributed to air leakage from the respiratory tract, is a common complication of interstitial lung disease often resulting in mediastinal widening. However, several cases of pneumomediastinum without mediastinal widening have been observed. This study aimed to investigate the cause of pneumomediastinum in patients without mediastinal widening.

**Patients and methods:** This study included 41 patients diagnosed with pneumomediastinum using computed tomography (CT) between July 2011 and September 2021 at Yokohama Minamikyosai Hospital; they had undergone a previous CT showing no gas density. Based on a comparison with previous CT images, the patients were classified into two groups: without mediastinal widening and with mediastinal widening.

**Results:** Of the 41 patients, 13 and 28 had pneumomediastinum without and with mediastinal widening, respectively. There were no significant differences in the sex, age, body mass index, or pneumomediastinum distribution between the two groups. However, the rate of weight loss per month was significantly higher in the group without mediastinal widening than in that with mediastinal widening. No significant differences were observed in the respiratory function test results between the two groups; however, 11 of the 13 patients had restrictive disorders. Pulmonary disease in this group included idiopathic pulmonary fibrosis (n = 6) and interstitial lung disease with collagen disease (n = 4). Pneumomediastinum occurred during periods of weight loss in all the patients excluding two patients without data.

**Conclusion:** Pneumomediastinum without mediastinal widening occurs during rapid weight loss and is often associated with restrictive lung disorders. The negative pressure attributed to the decreased plasticity of the lungs, which complements the space where the mediastinal fat has disappeared, is presumably the cause of pneumomediastinum. This pathophysiology is different from that of conventional pneumomediastinum attributed to increased intrapleural space pressure; thus, we propose to name the abovementioned pathophysiology negative pressure pneumomediastinum.

## Introduction

Pneumomediastinum (PM), which is a clinical condition indicated by the presence of gas in the mediastinum, is classified into spontaneous pneumomediastinum (SPM) and secondary PM according to the underlying etiology [1]. The pathophysiology of SPM is inferred based on a three-step process that identifies alveolar ruptures, air dissection along bronchovascular sheaths, and the spreading of pulmonary interstitial emphysema into the mediastinum [2]. The causes of secondary PM include esophageal injury, tracheal or bronchial injury related to trauma or of iatrogenic origin, and aerogenic infection.

SPM is a well-known complication of various respiratory diseases, including interstitial lung disease (ILD). In some cases, upon the progression of respiratory failure, mediastinal enlargement results from air leaks owing to an increase in the intrapleural pressure, and is often associated with pneumothorax. However, a case of a patient with ILD and PM without expansion of the mediastinal width has been reported previously [3]. In this case, the mediastinal fat decreased as a result of weight loss, and gas density appeared to replace the space. We presume that there is a decrease in the mediastinal pressure since the space left following mediastinal fat loss is not substituted by lung expansion owing to low compliance of the lung in patients with ILD. A decrease in the mediastinal pressure may result in the drawing of air from the airway or elution of nitrogen from the tissues, leading to a vacuum phenomenon, in which gas accumulates in the joint spaces or intervertebral discs. No pneumothorax or subcutaneous emphysema was observed in these cases since they were not caused by air leakage.

We hypothesized that this new pathophysiology should be named “negative pressure PM,” the pathologic characteristic of which is the decrease in the mediastinal pressure. This is distinct from conventional PM, which results from air leakage due to an increase in intrapleural pressure. Thus, the purpose of this study was to further investigate negative pressure PM and elucidate its etiology and clinical presentation.

## Materials and methods

This study was approved by the institutional review board of Yokohama Minamikyosai Hospital (29-1-10), which waived the need for informed consent owing to the retrospective nature of the study.

We reviewed a database of radiologically diagnosed cases of PM on computed tomography (CT) between July 2011 and September 2021 at Yokohama Minamikyosai Hospital. Of these, upon reviewing the images and clinical records, we excluded the patients without previously recorded CT studies for comparison of the mediastinum volume and those with an iatrogenic or traumatic PM; thus, 41 patients were included in the study.

CT images were obtained using a multidetector-row CT (Aquilion, Toshiba, Nasu Japan); images of the lung and mediastinal windows of 5-mm thickness were evaluated. We reviewed the images and identified PM and its distribution. In cases where the mediastinal contour in all the slices did not deviate outward compared with the previous CT images, no mediastinal enlargement was considered, which was classified as group A. Other cases were classified into group B since the mediastinum was presumed to have widened. The sites of the PM were the anterior mediastinum, which was ventral to the anterior margin of the ascending aorta; the posterior mediastinum, which was posterior to the dorsal margin of the trachea; the middle mediastinum, which was between the anterior and posterior mediastinum; the superior mediastinum, which was cranial to the aortic arch; and the hilum, which was the part beyond the lateral side of the mediastinum. Emphysema, which extends to the cranial side rather than the apex of the lung, is not considered mediastinal, but subcutaneous emphysema. Pneumothorax and subcutaneous emphysema were recorded.

We established a diagnosis of respiratory disease based on the patient’s medical history. In order to estimate body weight loss from two measurements of body weight taken around the time of the present CT, we calculated the rate of weight loss per month using the following equation:

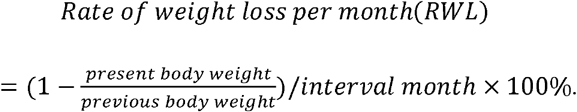

The presence or absence of chest pain and other symptoms was confirmed. We recorded the results of the respiratory function test performed at the time of the CT examination.

Statistical comparisons of age, body mass index (BMI), RWL, predicted vital capacity (%VC), FEV1.0% were performed using paired-samples t-tests; sex, pneumothorax, and subcutaneous emphysema with chi-square test; and the distribution of PM with chi-square test for goodness of fit using SPSS v. 21.0 for Windows (IBM, Armonk, NY, USA). Statistical significance was defined as *p* < 0.05.

## Results

Of the included 41 patients, 13 patients (9 male, 4 female) had no mediastinal widening, and 28 patients (21 males, 7 females) had mediastinal widening in the CT images as compared to the previous CT images (Table 1). The average age of the 13 patients was 71.8 (range: 29–90) years, while that of the 28 patients was 76.9 (range: 30–87) years. There were no significant differences in the patient group attributes.

**Table 1.**
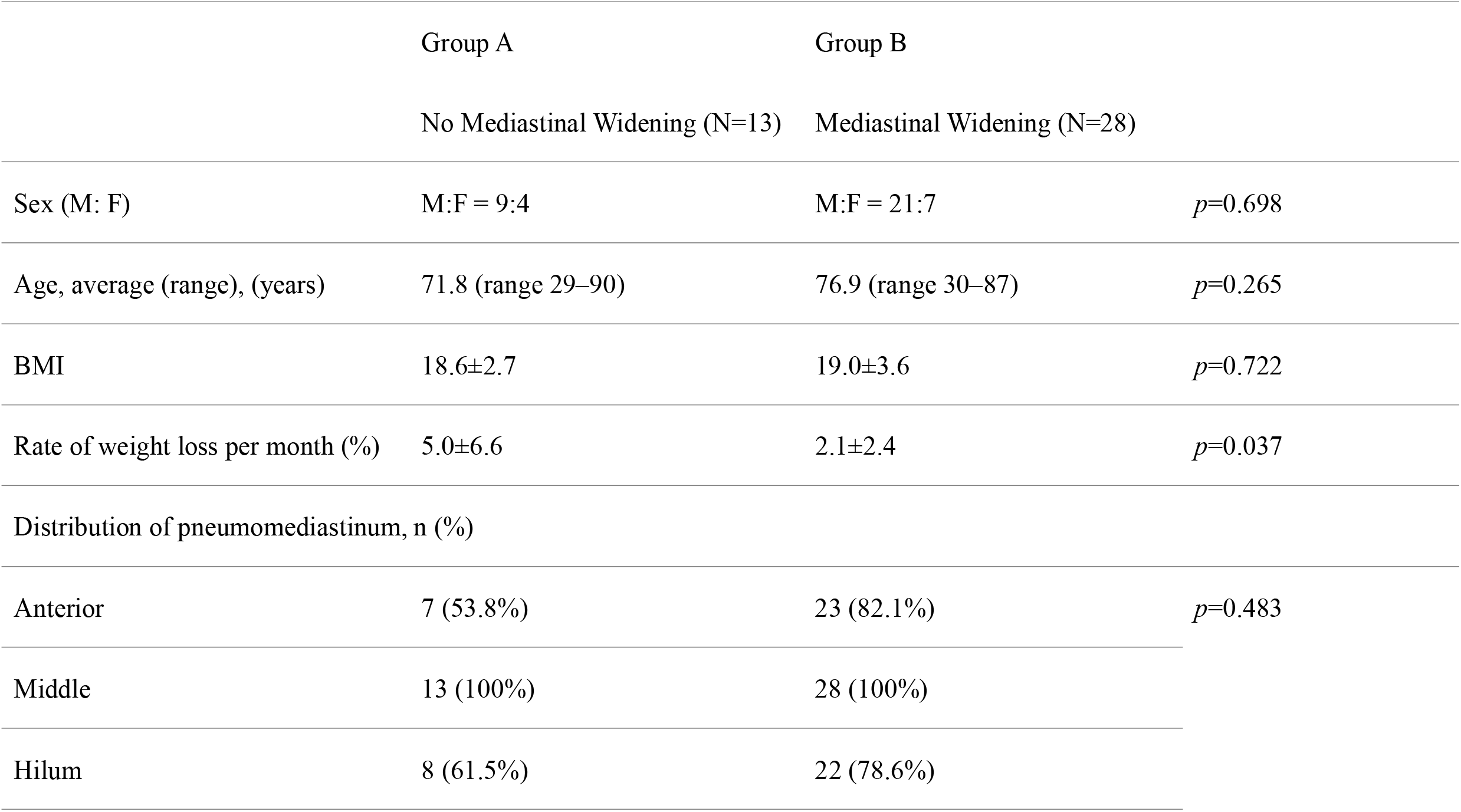
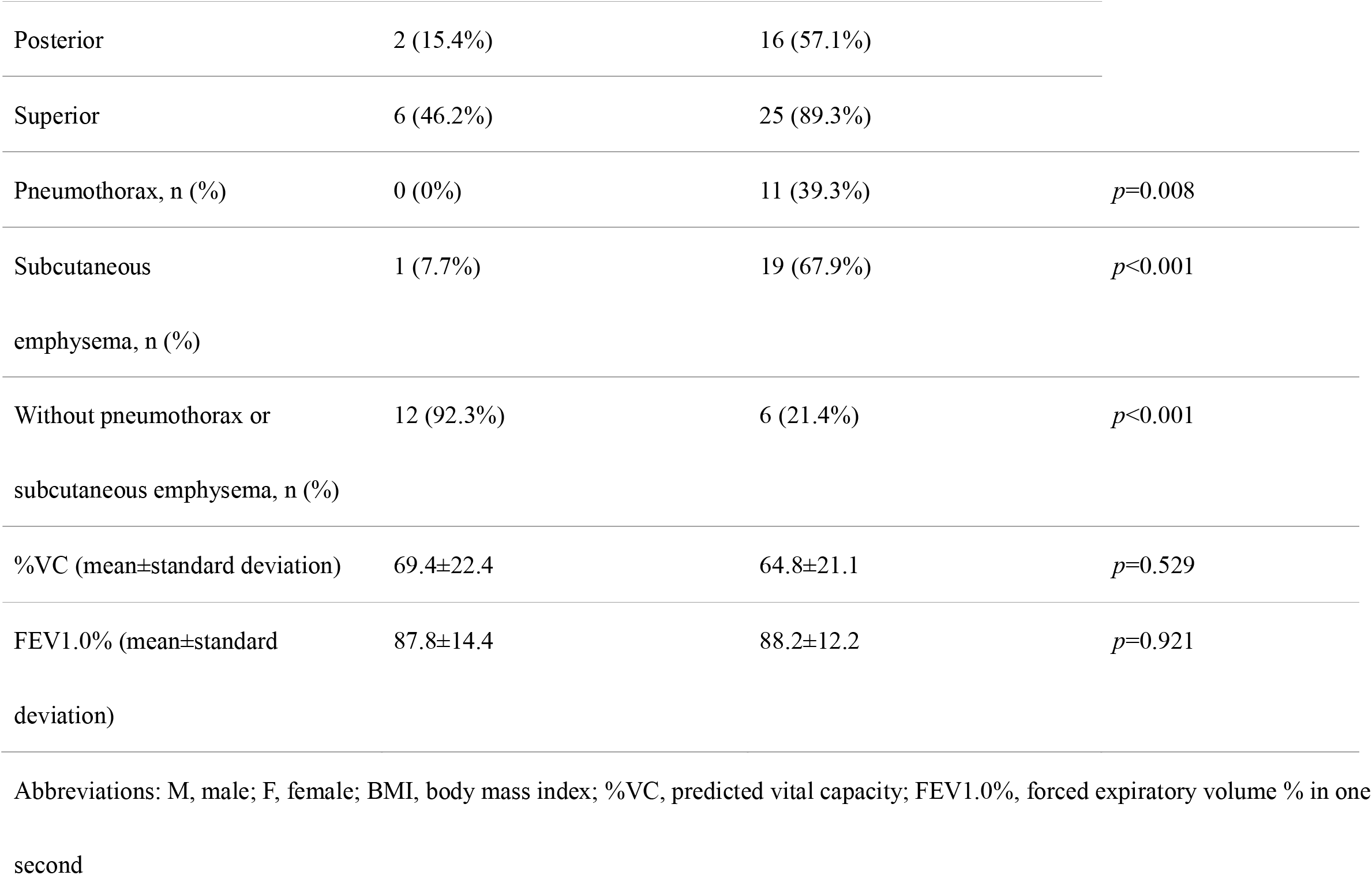
Patient characteristics.

The average BMI of group A and B were 18.6±2.7 and 19.0±3.6, without significant difference between the two groups. However, the average RWL of group A and B were 5.0±6.6 and 2.1±2.4, respectively. The rate of weight loss was significantly higher in the group without mediastinal widening than in the group with widening. There was no significant difference in the distribution of PM between the two groups.

Patients without mediastinal widening had subcutaneous emphysema or pneumothorax, except for one patient (case 8).

The diagnoses of pulmonary disease in these 13 patients included idiopathic pulmonary fibrosis (IPF, n =6), ILD with collagen disease (n =4), and other diseases (Table 2).

**Table 2.** Patient characteristics of the group with no mediastinal widening.

| Case | Age | Sex | Clinical diagnosis | Symptom | BMI | RWL | Distribution of gas |  |  |  |  | %VC | FEV1.0% |
| --- | --- | --- | --- | --- | --- | --- | --- | --- | --- | --- | --- | --- | --- |
|  |  |  |  |  |  |  | ant | mid | hilum | post | sup |  |  |
| 1 | In their 70s | F | SLE, ILD | Dyspnea | 18.6 | 19.6 | + | + | - | - | - | 66.5 | 69.39 |
| 2 | In their 70s | M | MCTD, ILD | No | 16.7 | 0.2 | - | + | - | - | - | 65.5 | 92.37 |
| 3 | In their 70s | M | IPF | No | 17.2 | 16.1 | - | + | - | - | + | 87.1 | 89.66 |
| 4 | In their 80s | M | IPF | Sputum, fever | 15.2 | 1.7 | - | + | + | - | - | 55.9 | 92.11 |
| 5 | In their 80s | M | CEP | Dyspnea | 19.9 | 1.1 | + | + | - | - | - | 40.3 | 49.3 |
| 6 | In their 80s | M | IPF | Appetite loss | 22.9 | 2.1 | + | + | + | + | - | 63.4 | 96.32 |
| 7 | In their 70s | M | IPF | Dyspnea | 24.1 | NA | + | + | + | + | + | 60.4 | 85.29 |
| 8 | In their 70s | M | GC. | No | 16.2 | 3.6 | - | + | + | - | + | 105 | 80.3 |
| 9 | In their 20s | M | UC | Fever | 20.1 | 5.5 | - | + | - | - | - | 118 | 92.28 |
| 10 | In their 50s | F | SjS, ILD | Dyspnea | 17.6 | 2.6 | + | + | + | - | - | 76.7 | 95.45 |
| 11 | In their 80s | F | IPF | Cough, chest pain | 16.2 | 1.3 | + | + | + | - | + | 46.2 | 100 |
| 12 | In their 70s | F | PMDM, SjS, ILD | Dyspnea | 17.5 | 1.0 | - | + | + | - | + | 65.6 | 98.54 |
| 13 | In their 90s | M | IPF | Fever | 20.2 | NA | + | + | + | - | + | 52 | 100 |

Abbreviations: ant, anterior mediastinum;; BMI, body mass index; CEP, chronic eosinophilic pneumonia; GC, gastric cancer; ILD, interstitial lung disease; IPF, idiopathic pulmonary fibrosis; MCTD, mixed connective tissue disease; mid, middle mediastinum; N/A, not available; PMDM, dermatomyositis/polymyositis; post, posterior mediastinum; RWL, rate of weight loss per month; SjS, Sjogren syndrome; SLE, systemic lupus erythematosus; sup, superior mediastinum; UC, ulcerative colitis; %VC, predicted vital capacity

Among the 13 patients, except for two patients without data, the CT examination revealed gas densities in the mediastinum that corresponded to the time when they lost weight. There was no complaint of chest pain with the exception of one patient. The respiratory function test revealed a restrictive disorder with a %VC of < 80% in 13 patients except in two patients with gastric cancer and ulcerative colitis. There were no significant differences in the %VC and FEV1.0% between groups A and B.

## Representative cases

### Case 1

Case 1 included a woman in her 70s who was admitted to the hospital with a history of cough, dyspnea, and slight fever for three months. Her body temperature was 37.1 °C, and a fine crackle was detected in the dorsum on auscultation. Chest computed tomography (CT) revealed reticular opacities and consolidation, which suggested a nonspecific interstitial pneumonia (NSIP) pattern on the bases of both lungs (Fig 1A). She was diagnosed with systemic lupus erythematosus with Sjogren’s syndrome based on the presence of facial eruptions for several years, intraoral drying, antinuclear antibodies identified by blood tests, high levels of CH50, and anti-RNP antibodies. The patient was treated with prednisolone and mycophenolate mofetil. The dyspnea gradually improved. A CT scan performed one month later to evaluate the effect of NSIP treatment revealed PM (Fig 1B).

**Fig 1.**
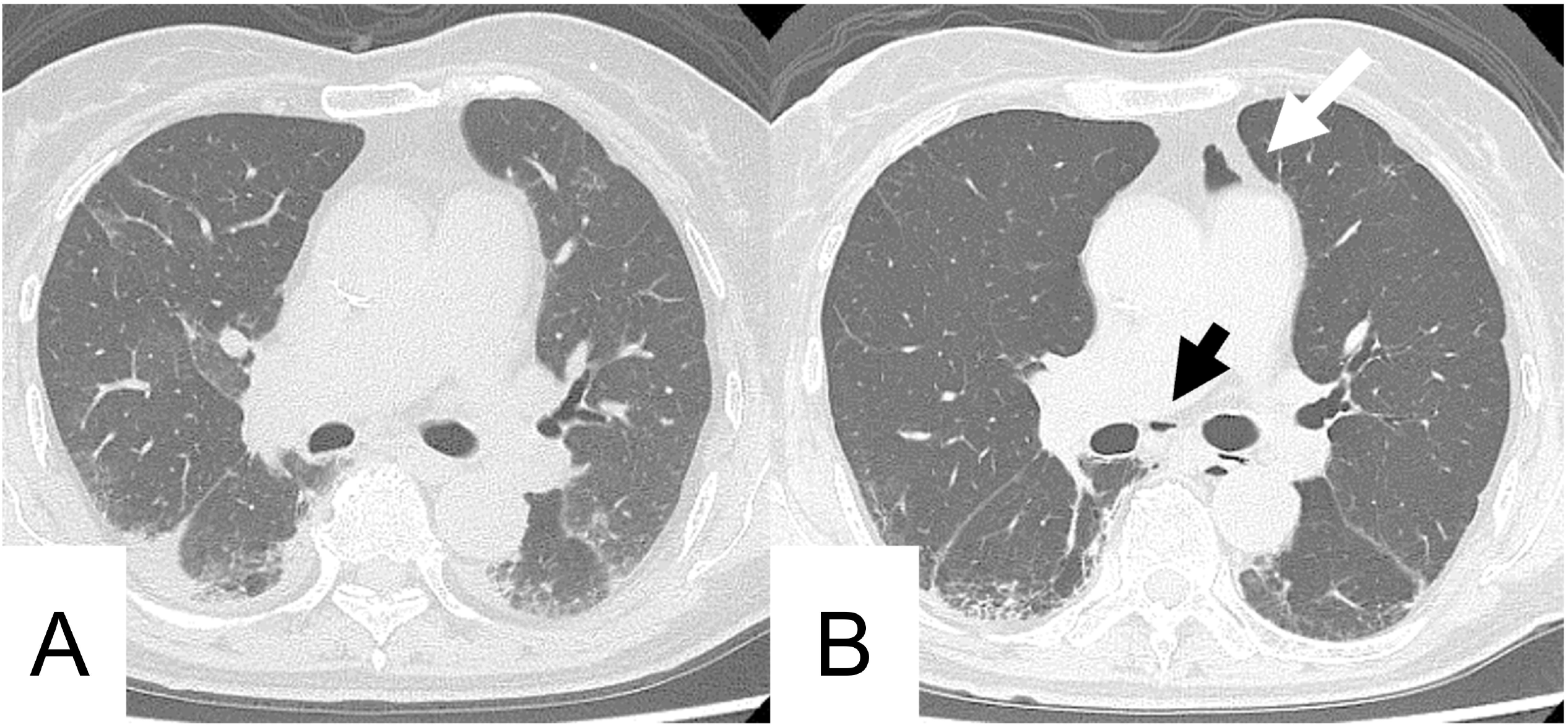
Computed tomography images of the chest of a woman in her 70s with systemic lupus erythematosus and Sjogren syndrome. Computed tomography (CT) performed at admission revealed consolidation and reticular opacities in peripheral areas of both the lungs (A). A CT scan performed one month later revealed the appearance of gas density in the anterior mediastinum (white arrow) and around the carina (black arrow) (B). The width of the anterior mediastinum decreased owing to weight loss.

### Case 4

Case 4 included a man in his 80s with idiopathic pulmonary fibrosis (IPF). Lung fibrosis was identified by CT one year prior to presentation (Fig 2A).

**Fig 2.**
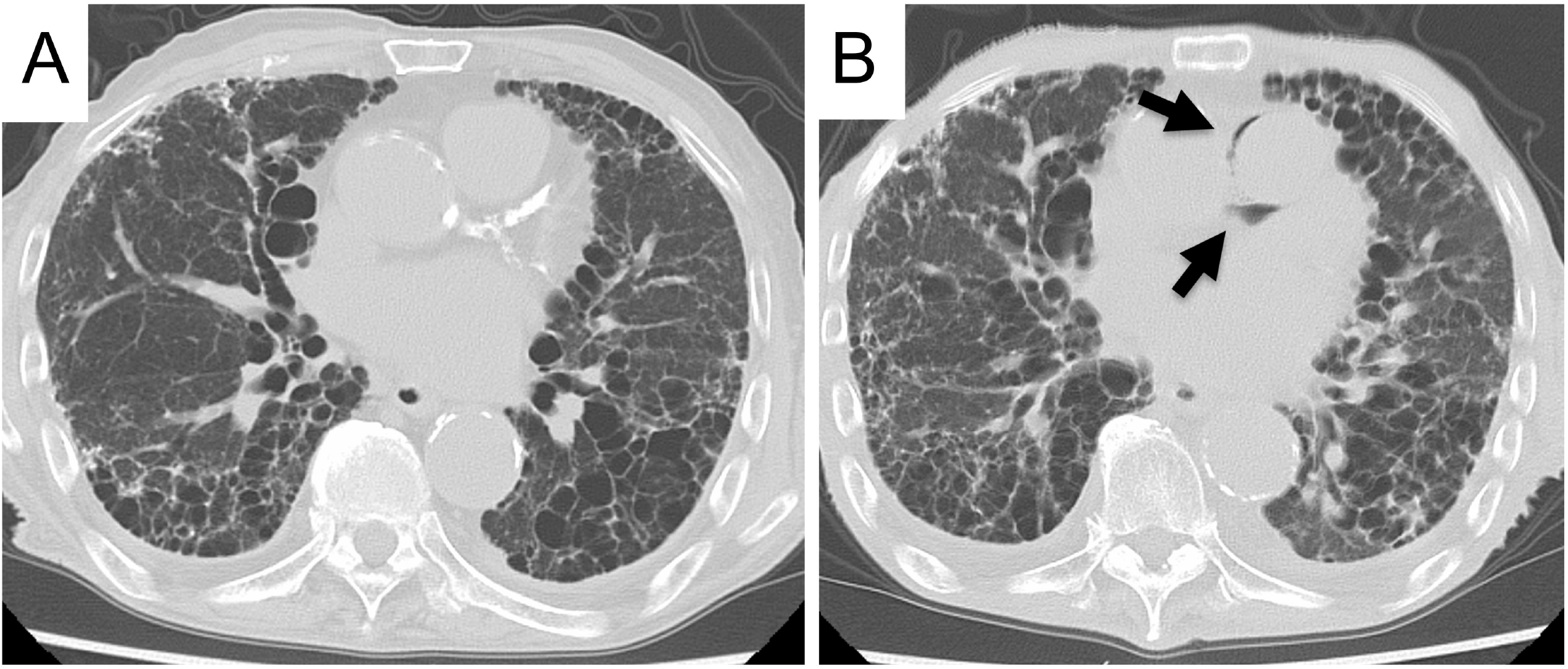
Computed tomography images of the chest of a man in his 80s with idiopathic pulmonary fibrosis. Computed tomography (CT) performed a year ago revealed reticular opacities and honeycombing in peripheral areas of both lungs consistent with IPF (A). A CT scan performed on admission revealed reticular opacities and honeycombing were exacerbated. PM appeared around the pulmonary artery (arrows); however, no enlargement of the mediastinum (B) was observed.

He was admitted to the hospital with a history of one week of productive cough, dyspnea, and fever. His body temperature was 38.2 °C, and a fine crackle was detected in the dorsum on auscultation. Chest CT revealed reticular opacities and honeycombing, suggesting a usual interstitial pneumonia pattern in both peripheral lungs, which were exacerbated from prior CT a year ago (Fig 2B). PM appeared around the pulmonary artery; however, no enlargement of the mediastinum was observed. Subcutaneous fat disappeared on the CT images, consistent with his weight loss from 43.5 kg to 33.0 kg over the past year.

## Discussion

SPM is presumed to be caused by the leakage of air from the respiratory tract. Our study aims to elucidate the reason for the absence of expansion of the mediastinum in certain cases despite the additional volume of increased air.

SPM is a well-known complication of various ILDs, including IPF and connective tissue disorders [4–6], with increased intrapleural pressure owing to cough and constrained respiration, which are thought to produce alveolar rupture. One report suggested that corticosteroids, which are administered to treat primary disease, damage the interstitial tissues of the lungs [7], and PM also frequently occurs when the condition worsens if steroids and immunosuppressive drugs are not administered [8]. These studies mainly focused on the clinical characteristics of lung diseases that lead to air leaks. However, our study focused on the relative pressure of the mediastinum and not on the cause of the air leaks. We hypothesized that PM is caused by the pressure gradient between the intrapleural space and mediastinum (Fig 3). The pressure in the intrapleural space and mediastinum is balanced in the normal chest; however, when the intrapleural pressure rises, this balance is disrupted, resulting in a pressure gradient, and an air leak is produced to balance the pressure. This conventional PM is referred to as positive pressure PM since it is attributed to the positive pressure in the intrapleural space. Similarly, negative pressure PM occurs to eliminate the pressure difference during decreased mediastinal pressure.

**Fig 3.**
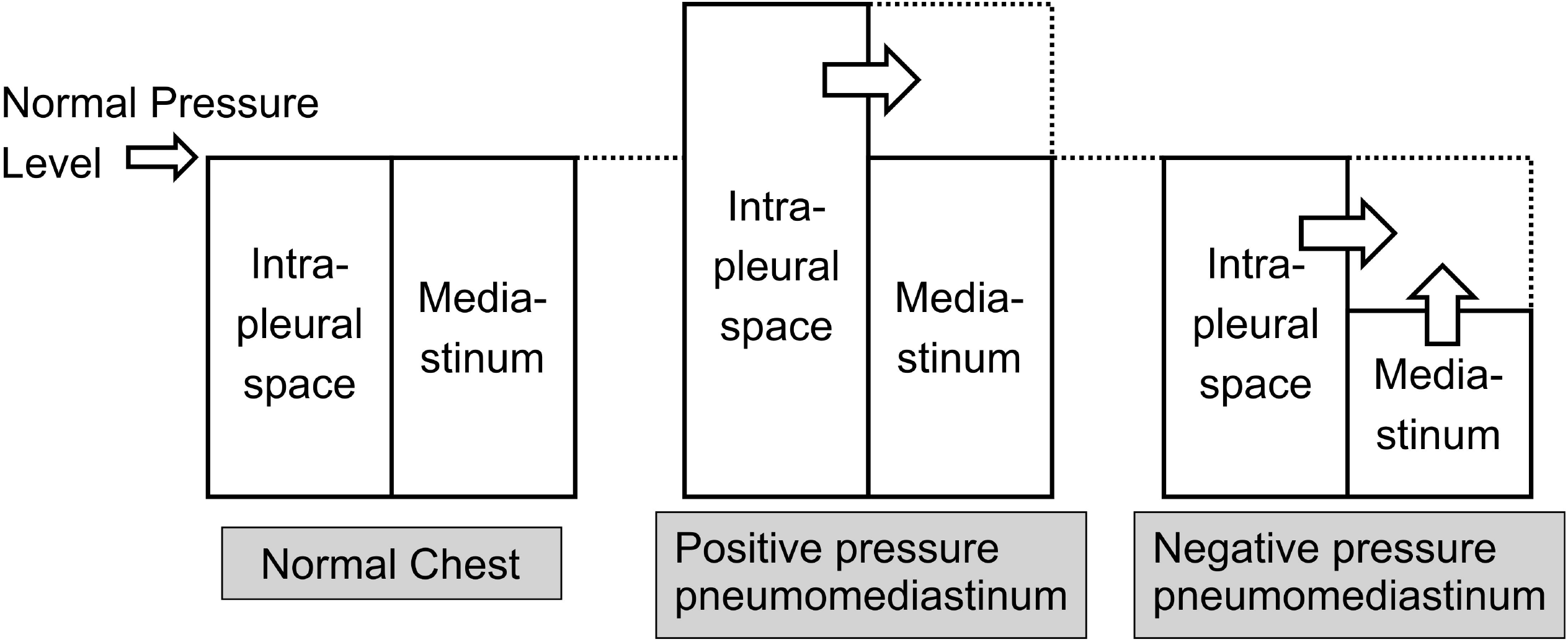
The balance between the pressures in the intrathoracic cavity and the mediastinum in normal chest. When the intrapleural space pressure rises for some reason, a positive pressure pneumomediastinum is caused to balance the difference. Similarly, negative pressure pneumomediastinum occurs to abolish the pressure difference when the mediastinal pressure decreases.

The conditions wherein the mediastinal pressure decreases are very limited. In positive pressure PM, air leaks from the respiratory tract increase the mediastinal pressure and mediastinal widening (Fig 4). When body weight loss and mediastinal fat are reduced in normal lungs, the capacity of the lungs increases and the space is replaced. As a result, the width of the mediastinum is narrowed and the pressure remains unaltered. However, in patients with restrictive lung disorders due to ILD, remodeling and replacement do not occur owing to the decreased plasticity of the lung. This reduces the mediastinal pressure and causes a negative pressure PM without mediastinal resizing. The faster rate of weight loss in Group A also supports this hypothesis.

**Fig 4.**
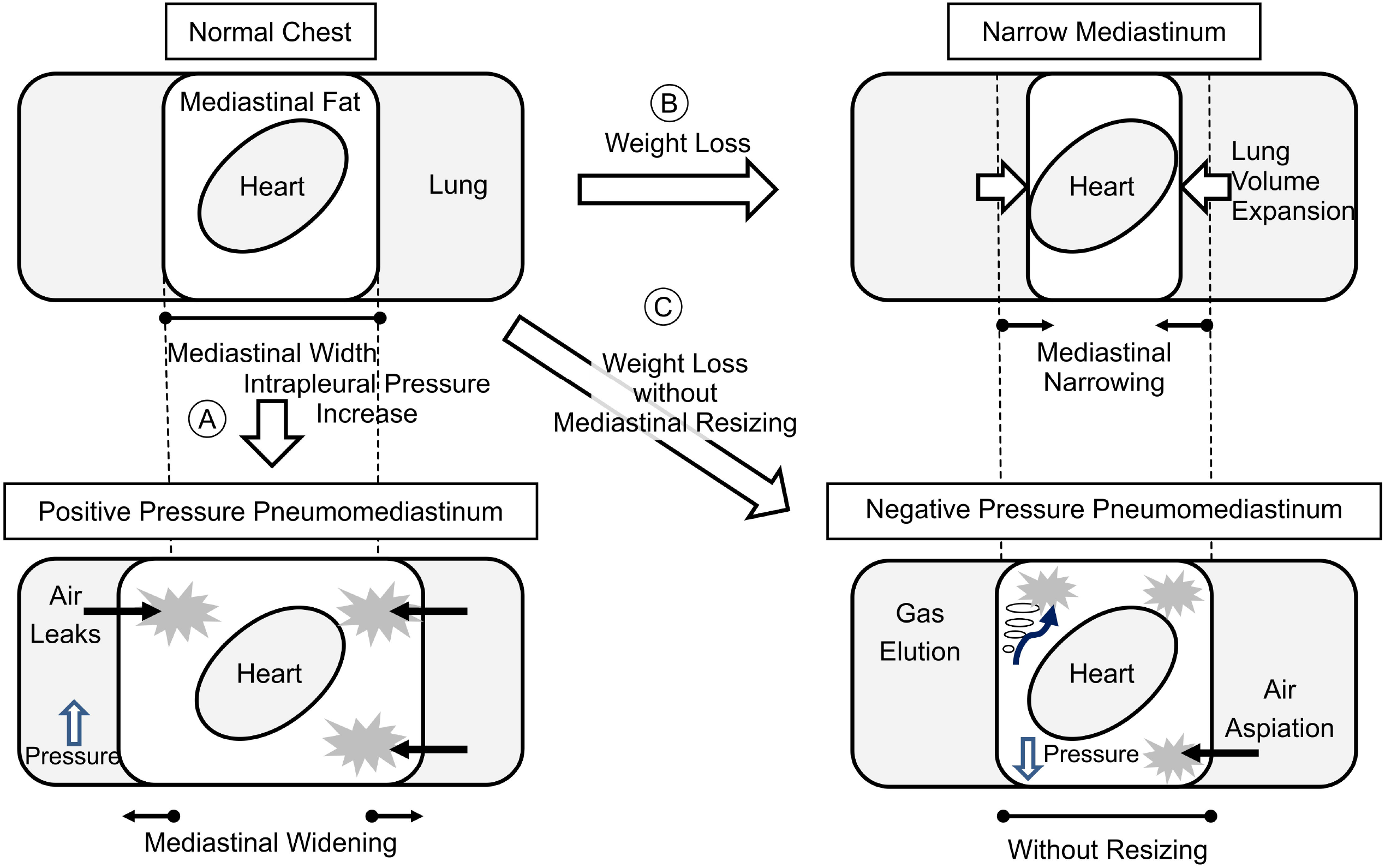
Pathophysiology of positive and negative pressure pneumomediastinum. Increased intrapleural pressure (A) causes air leaks, resulting in positive pressure pneumomediastinum. Mediastinal fat decreases with weight loss (B) resulting in mediastinal narrowing because the lungs replace the space. In patients with restrictive lung disorder due to interstitial lung disease, mediastinal resizing does not occur owing to reduced plasticity. This reduces the mediastinal pressure and causes pneumomediastinum without mediastinal resizing (C).

There are two possible reasons for gas collection. One is by air from the intrathoracic airway, similar to conventional SPM, and the other is by the vacuum phenomenon (VP). VP is a state in which gas is retained in the synovial joints or intervertebral discs. VP is most frequently associated with degenerative joint diseases but has also been associated with other pathologies, such as osteonecrosis and compression fracture; therefore, its etiology is still under discussion [9–12]. The gas within the intervertebral discs contains at least 90% nitrogen combined with oxygen, carbon dioxide, and other trace gases [13]. Unlike oxygen, nitrogen collected in the dead space is not metabolized and is thus thought to accumulate in the tissue. The VP adequately explains the gas collection isolated from the airways and hilum.

In the past, all the cases of PM of unknown cause were classified as SPM. However, our results show evidence of the existence of negative-pressure PM; therefore, we propose that SPM should be classified into two categories (Fig 5): new additional category of negative pressure PM and positive pressure PM (which represents conventional SPM). During lung disease treatment interventions, PM should be distinguished from positive pressure PM attributed to exacerbation of lung disease and negative pressure PM attributed to weight loss.

**Fig 5.**
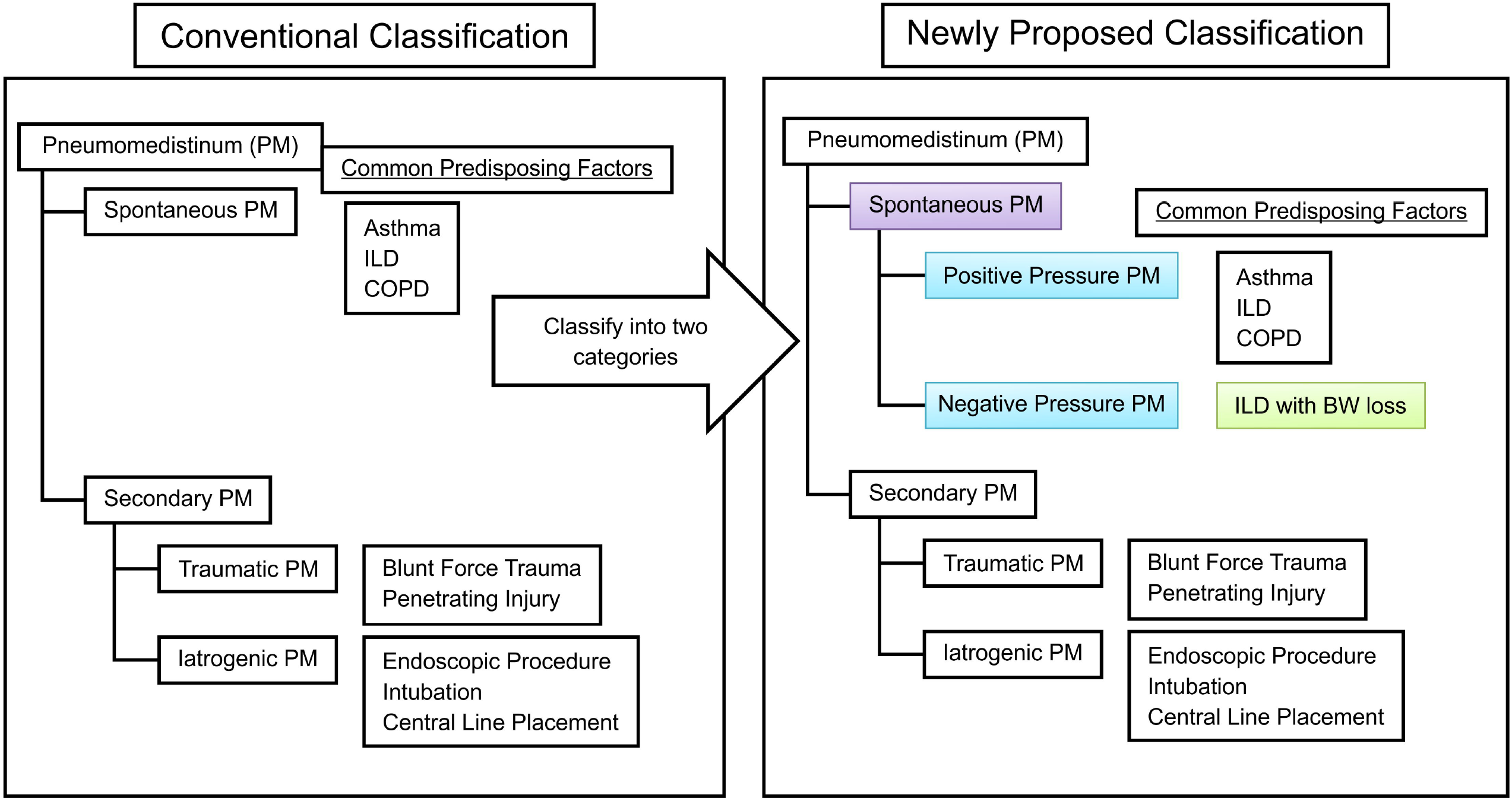
Newly proposed classification for spontaneous pneumomediastinum. Pneumomediastinum (PM) is classified as spontaneous PM and secondary PM as previously mentioned. Spontaneous PM can be divided into two categories: new additional category of negative pressure PM and positive pressure PM that represents conventional spontaneous PM. BW, body weight; COPD, chronic obstructive pulmonary disease; ILD, interstitial lung disease; PM, pneumomediastinum.

This study has certain limitations. First, this was a retrospective study, and CT scans and body weight measurements were not performed simultaneously; thus, the precise rate of body weight loss could not be calculated. However, these measurements allowed us to estimate the weight loss based on the decrease in the subcutaneous fat and mediastinal fat compared to that in the initial CT before identification of gas density. Similarly, the respiratory function test did not reflect the respiratory function at the precise time of the CT. Second, the presence or absence of mediastinal enlargement was manually judged by a radiologist; therefore, accurate classification was not possible in cases with a small amount of PM. Third, sampling and component analysis of gas that accumulated in the mediastinum were not performed. These tests can be used to determine whether the accumulated gas originates from the respiratory tract or is caused by the VP.

## Conclusion

In a certain number of cases of PM included in this study, the mediastinum was not enlarged. These were not caused by air leaks owing to an increase in the intrapleural pressure, but by a decrease in the mediastinal pressure owing to mediastinal fat volume loss combined with lung plasticity reduction in ILD. Since the cause of PM is presumed to be associated with the disruption of the relative pressure balance between the intrapleural space and mediastinum, SPM can be divided into positive pressure PM, which increases intrapleural space pressure, and negative pressure PM, which decreases mediastinal pressure.

## Data Availability

All relevant data are available from the Open Science Framework (https://osf.io/https://osf.io/vzds9/).

## Acknowledgments

We would like to thank Dr. T. Iwasawa for useful discussions. We would like to thank Editage (www.editage.jp) for English language editing.

## Notes

### Competing Interest Statement

The authors have declared no competing interest.

### Funding Statement

The author(s) received no specific funding for this work.

## References

1. Sahni S, Verma S, Grullon J, Esquire A, Patel P, Talwar A. Spontaneous pneumomediastinum: time for consensus. N Am J Med Sci. 2013;5: 460–464. doi: 10.4103/1947-2714.117296.

2. Macklin MT, Macklin CC. Malignant interstitial emphysema of the lungs and mediastinum as an important occult complication in many respiratory diseases and other conditions: an interpretation of the clinical literature in the light of laboratory experiment. Medicine. 1944;23: 281–358. doi: 10.1097/00005792-194412000-00001.

3. Hagiwara H, Torii I. Mediastinal vacuum phenomenon: atypical pneumomediastinum caused by gas replacement of diminished fat. Int Med Case Rep J. 2015;8: 283–286. doi: 10.2147/IMCRJ.S93664.

4. Fujiwara T. Pneumomediastinum in pulmonary fibrosis. Detection by computed tomography. Chest. 1993;104: 44–46. doi: 10.1378/chest.104.1.44.

5. Kono H, Inokuma S, Nakayama H, Suzuki M. Pneumomediastinum in dermatomyositis: association with cutaneous vasculopathy. Ann Rheum Dis. 2000;59: 372–376. doi: 10.1136/ard.59.5.372.

6. Franquet T, Giménez A, Torrubia S, Sabaté JM, Rodriguez-Arias JM. Spontaneous pneumothorax and pneumomediastinum in IPF. Eur Radiol. 2000;10: 108–113. doi: 10.1007/s003300050014.

7. Yamanishi Y, Maeda H, Konishi F, Hiyama K, Yamana S, Ishioka S, et al. Dermatomyositis associated with rapidly progressive fatal interstitial pneumonitis and pneumomediastinum. Scand J Rheumatol. 1999;28: 58–61. doi: 10.1080/03009749950155805.

8. Matsuoka S, Kurihara Y, Yagihashi K, Okamoto K, Niimi H, Nakajima Y. Thin-section CT assessment of spontaneous pneumomediastinum in interstitial lung disease: correlation with serial changes in lung parenchymal abnormalities. Respir Med. 2006;100: 11–19. doi: 10.1016/j.rmed.2005.04.016.

9. Resnick D, Niwayama G, Guerra J Jr, Vint V, Usselman J. Spinal vacuum phenomena: anatomical study and review. Radiology. 1981;139: 341–348. doi: 10.1148/radiology.139.2.7220878.

10. Libicher M, Appelt A, Berger I, Baier M, Meeder PJ, Grafe I, et al. The intravertebral vacuum phenomen as specific sign of osteonecrosis in vertebral compression fractures: results from a radiological and histological study. Eur Radiol. 2007;17: 2248–2252. doi: 10.1007/s00330-007-0684-0.

11. Lafforgue P, Chagnaud C, Daumen-Legré V, Daver L, Kasbarian M, Acquaviva PC. The intravertebral vacuum phenomenon (“vertebral osteonecrosis”). Migration of intradiscal gas in a fractured vertebral body? Spine (Phila Pa 1976). 1997;22: 1885–1891. doi: 10.1097/00007632-199708150-00015.

12. Patten RM. Vacuum phenomenon: a potential pitfall in the interpretation of gradient-recalled-echo MR images of the shoulder. AJR Am J Roentgenol. 1994;162: 1383–1386. doi: 10.2214/ajr.162.6.8192004.

13. Ford LT, Gilula LA, Murphy WA, Gado M. Analysis of gas in vacuum lumbar disc. AJR Am J Roentgenol. 1977;128: 1056–1057. doi: 10.2214/ajr.128.6.1056.

